# The sound of snoring: AI-based acoustic phenotyping of upper airway obstruction in Obstructive Sleep Apnea

**DOI:** 10.64898/2026.09.14.26362626

**Authors:** Federico Leone, Francesco Pietrogiacomi, Emanuele Agrimi, Linda Fiorini, Federica Vultaggio, Alessandro Bianchi, Valerio Cesarini, Giovanni Costantini, Giorgio Gnecco, Fabrizio Salamanca, Francesco Mozzanica

**Affiliations:** Department of Otorhinolaryngology, Sleep Surgery Center, Sleep Disorders Center, Istituto Auxologico Italiano IRCCS, Milan, Italy; Department of Clinical Sciences and Community Health, Dipartimento di Eccellenza 2023-2027, Universita degli Studi di Milano, Milan, Italy; IMT School for Advanced Studies Lucca, Lucca, Italy; University of Rome Tor Vergata, Department of Electronic Engineering, Rome, Italy; Department of Otorhinolaryngology, IRCCS Multimedica, Milan, Italy

## Abstract

**Background:** Drug-Induced Sleep Endoscopy (DISE) is the current reference standard for identifying the anatomical site of upper airway obstruction in obstructive sleep apnea (OSA), but its invasiveness and limited availability restrict its routine use. Because snoring is generated by vibration of the obstructing upper-airway structures, its acoustic characteristics may provide a non-invasive biomarker of the anatomical site of obstruction. This study investigated whether machine learning could reliably distinguish palatal from epiglottic snoring using acoustic information alone.

**Methods:** A retrospective analysis was performed on 159 DISE recordings obtained from adult patients with moderate, non-positional OSA. Snoring events were independently identified and anatomically classified by two blinded expert examiners. Only events with complete inter-observer agreement were included. The corresponding audio segments were extracted and characterized using spectral, cepstral, temporal and harmonic acoustic features. Feature selection was performed using recursive feature elimination. Support Vector Machine (SVM) and Multilayer Perceptron (MLP) classifiers were developed using a patient-independent nested cross-validation framework.

**Results:** The final dataset comprised 1,759 snoring events, including 1,054 palatal and 705 epiglottic recordings. Both classifiers demonstrated robust discrimination between the two anatomical classes under patient-independent validation, achieving ROC AUC values of 0.90± 0.03 (SVM) and 0.90± 0.03 (MLP), with balanced accuracies of 0.83 and 0.81, respectively. The most informative predictors were spectral features, particularly spectral energy distribution, spectral flux, Mel-Frequency Cepstral Coefficients (MFCCs), and spectral flatness, whereas fundamental frequency contributed minimally to classification.

**Conclusions:** Snoring contains reproducible acoustic information reflecting the anatomical origin of upper-airway obstruction. By combining high-confidence DISE-derived anatomical labels with a rigorous machine learning framework, this study demonstrates the feasibility of non-invasive acoustic phenotyping of clinically relevant obstruction sites. Rather than proposing a novel artificial intelligence algorithm, our work establishes a clinically oriented framework that may support future decision-support tools for patient selection, treatment planning, and multicentre development of comprehensive acoustic phenotyping models.

## Introduction

Obstructive Sleep Apnea (OSA) is one of the most common sleep-related breathing disorders, characterized by recurrent episodes of upper airway narrowing or collapse during sleep. These events result in reduced (hypopnea) or absent (apnea) airflow, intermittent hypoxia, vibration of upper airway tissues (snoring) and recurrent arousals (sleep fragmentation) (1).

OSA is highly prevalent yet frequently underrecognized. Mild OSA, conventionally defined by an apnea-hypopnea index of 5-15 events per hour of sleep, is estimated to affect approximately 10-25% of the general adult population worldwide (1,2). Common clinical manifestations include habitual snoring, disrupted sleep, witnessed apneas. Moreover, OSAS is associated with excessive daytime sleepiness, reduction of quality of life (QOL), and increased incidence of motor vehicle accidents. In addition, it represents an independent risk factor for several adverse cardiovascular outcomes, such as arterial hypertension, ischemic heart disease, arrhythmias, and ischemic stroke (1–6). Thus, early identification and appropriate treatment are therefore essential.

Polysomnography represents the reference standard for the diagnosis of OSA, while continuous positive airway pressure (CPAP) is considered the first-line treatment for moderate-to-severe disease (7). However, suboptimal long-term adherence is frequently reported and this might limit its effectiveness in clinical practice (7). Consequently, several alternative or complementary therapeutic strategies have become integral to contemporary OSA management, including upper airway surgery, mandibular advancement devices (MAD), positional therapy, myofunctional therapy, and hypoglossal nerve stimulation (8–13).

Each of the above-mentioned approach has specific indications and appropriate treatment selection requires information about the anatomical site and pattern of upper airway narrowing and collapse. In this context, Drug-Induced Sleep Endoscopy (DISE) represents a valuable diagnostic tool able to provides direct visualization of upper airway dynamic under pharmacologically induced sleep-like conditions. It is currently regarded as the reference procedure for identifying obstruction sites and patterns and for guiding individualized treatment decisions (14). Nevertheless, DISE remains largely confined to selected specialized referral centers because it requires dedicated equipment, anesthesiologic support, trained personnel, and an appropriate clinical setting (14–16).

The development of rapid, minimally invasive, and widely accessible methods capable of providing clinically meaningful information on upper airway dynamics during sleep therefore represents a major unmet need in sleep medicine.

Snoring represents a particularly attractive candidate for this purpose. Rather than merely representing a symptom of airway obstruction, snoring is the acoustic consequence of tissue vibration generated by airflow through a narrowed and collapsible upper airway. Since the acoustic properties of a sound depend on the physical characteristics and anatomical location of its vibrating source, different sites and patterns of upper airway obstruction may produce distinct acoustic signatures (17). Although such differences may not be reliably distinguishable by human perception, they could potentially be detected through quantitative analysis of the acoustic signal.

Recent advances in artificial intelligence (AI), particularly machine learning (ML) (a subset of AI that focuses on training computer systems to learn and improve automatically from experience, without explicit programming) have enabled the identification of subtle and highly complex patterns within biomedical signals that may remain undetectable using conventional analytical methods. By learning relationships between patient-derived data and clinically relevant outcomes, AI and ML algorithms may support diagnostic classification, outcome prediction, and individualized treatment selection (18,19). Although these approaches have shown promising applications in OSA screening and severity estimation, considerably less attention has been devoted to the anatomical characterization of upper airway obstruction. Furthermore, the available evidence is limited by relatively small datasets, heterogeneous methodologies, and the frequent absence of clinically validated reference standards, hindering translation into routine clinical practice (20–22).

Therefore, the aim of this study was to investigate whether ML algorithms could identify the anatomical site of upper airway obstruction using only the acoustic characteristics of snoring events. Specifically, we developed and validated predictive models in order to provide proof of concept for snoring as a non-invasive acoustic biomarker of upper airway obstruction.

## Materials and Methods

### Dataset generation

This retrospective study was conducted at a tertiary referral center using anonymized drug-induced sleep endoscopy (DISE) recordings from adult patients with obstructive sleep apnea (OSA). All examinations had been performed by the same experienced sleep surgeon. The recordings were reviewed to generate a dataset of anatomically labelled audio tracks for the development and evaluation of machine-learning algorithms.

DISE recordings were eligible for inclusion according to the following criteria:

- adequate visualization of the upper airway from the velum to the epiglottis;
- absence of motion artifacts or incomplete visualization of the relevant anatomical structures;
- availability of synchronized, high-quality audio suitable for acoustic analysis;
- normal body weight, defined as a body mass index (BMI) <25 kg/m^2^;
- moderate, non-positional OSA, defined as a respiratory event index (REI) between 15 and 30 events/hour and a supine-to-lateral apnea-hypopnea index (AHI) variability <20% on full-night polygraphy.

Each eligible DISE recording was independently reviewed by two expert examiners who were blinded to each other’s assessments. Using the synchronized endoscopic video and audio, both examiners manually identified all snoring events and independently assigned each event to the corresponding anatomical site. Only events identified by both examiners and assigned to the same anatomical category were retained. For each concordant event, the corresponding synchronized video-audio segment was extracted as an individual clip in AVI format and labelled as palatal or epiglottic. This consensus-based procedure provided a high-confidence, DISE-derived anatomical ground truth for the subsequent machine-learning analysis.

The corresponding audio track was then extracted from each annotated video clip and manually trimmed to remove silent intervals and background noise using Reaper (23). The resulting audio files were exported as mono-channel PCM WAV recordings with a sampling rate of 44.1 kHz and a 16-bit resolution, constituting the dataset used for subsequent preprocessing and machine learning analysis.

The study was approved by the local Ethics Committee (Protocol No. 4150-ID), and all procedures were conducted in accordance with the Declaration of Helsinki.

### Data processing

Following extraction and trimming, the audio samples underwent volume normalization using the Root Mean Square (RMS) normalization to account for variations in microphone distance, sensitivity, and patient movement. The RMS value of a discrete signal *x[n]* of length *N* was computed for each audio clip as:

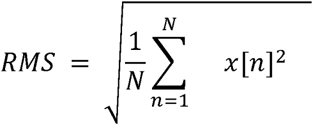

The audio samples were then scaled so that each clip had the same RMS value, using the lowest as a target in order to avoid positive gain that could lead to clipping, in turn ensuring that the distribution of signal amplitudes was standardized across all recordings. Finally, all the clips were resampled from 44,100 Hz to 22,050 Hz to optimize computational efficiency and ensure a uniform frequency range for subsequent feature extraction. This choice was made to reduce the number of samples for computational efficiency while taking into account that snoring-like sounds contain most of their relevant information at lower frequencies (24). Moreover, the selected down-sampling rate still preserves frequency content until 10 kHz. These preprocessing steps ensured the reliability and comparability of the acoustic data for the classification tasks.

### Feature Extraction

A comprehensive set of acoustic descriptors was extracted from each preprocessed snoring segment to characterize its cepstral, spectral, temporal, and harmonic properties. The cepstral descriptors included: Mel-Frequency Cepstral Coefficients (MFCCs) (25) and their first- and second-order derivatives. The spectral descriptors included: spectral flatness (26), spectral centroid (19), spectral bandwidth (27), spectral roll-off (28), spectral contrast, chroma features (29), spectral flux (30), energy distribution, and formants. The temporal descriptors included: zero-crossing rate and autocorrelation-based measures. Additional descriptors included: Harmonics-to-Noise Ratio (HNR), Cepstral Peak Prominence (CPP), estimated Vocal Tract Length (VTL), fundamental frequency (F0), and voicing ratio.

This process generated 691 acoustic features for each snoring event. Particular emphasis was placed on spectral descriptors because of their established relevance in snoring and biomedical sound analysis. F0-related measures were also included to investigate whether palatal and epiglottic snoring exhibited different periodic vibration characteristics despite the predominantly noise-like nature of snoring. A complete description of the extracted features, computational definitions, and implementation details is provided in the Supplementary Materials Appendix A.

### Classification, feature selection, and model validation

Two machine-learning models—a Support Vector Machine (SVM) and a Multi-Layer Perceptron (MLP)—were developed to classify snoring events as palatal or epiglottic since these two anatomical phenotypes represent the principal surgical targets in contemporary sleep surgery. Before model training, highly correlated acoustic features were removed, and recursive feature elimination using a linear SVM was applied to identify a final subset of 20 informative descriptors.

Model development and evaluation were performed using 5×5 nested cross-validation. The inner loop was used for feature selection and hyperparameter optimization, whereas the outer loop provided an unbiased estimate of model performance on held-out data. Standardization, feature selection, class balancing, and model optimization were performed independently within each training fold, without access to the corresponding validation or test data. All dataset partitions were performed at the patient level, ensuring that snoring events from the same patient could not appear in different data subsets, while class stratification was maintained whenever compatible with patient-wise splitting. Class imbalance was addressed using subject- and class-based sample weighting for the SVM and random oversampling for the MLP. Performance was evaluated using balanced accuracy as the primary metric, together with precision, recall, F1-score, and the area under the receiver operating characteristic curve (AUC-ROC). Full details of feature selection, hyperparameter optimization, class-balancing procedures, and fold composition are provided in the Supplementary Materials.

## Results

Following manual annotation and preprocessing, the final dataset comprised 1,759 audio segments obtained from 159 DISE recordings. Of these, 1,054 were classified as palatal snoring and 705 as epiglottic snoring. The average number of segments per subject was 11.1, with a mean segment duration of 1.28 s.

### Model performance on held-out test sets

The classification performance across the five test folds is summarized in the confusion matrices (Figure 1), and the metrics, reported as the mean values aggregated across the five folds, are shown in Table 2 and Table 3. Overall, the SVM achieved slightly better performance with a balanced accuracy of 0.83 and a ROC AUC of 0.90, compared to the MLP’s 0.81 and 0.90, respectively. This better performance was consistently reflected across class-specific metrics: the SVM achieved a precision of 0.85 for palatal events and 0.79 for epiglottic samples (with F1-scores of 0.85 and 0.78), while the MLP reached a precision of 0.85 and 0.75 (with F1-scores of 0.83 and 0.76). In both models, performance was notably higher for the palate class than for the epiglottis class, a trend likely driven by the inherent class imbalance. These results indicate that while both models effectively capture relevant anatomical information, the SVM provides marginally better classification effectiveness across all measured parameters.

**Figure 1.**
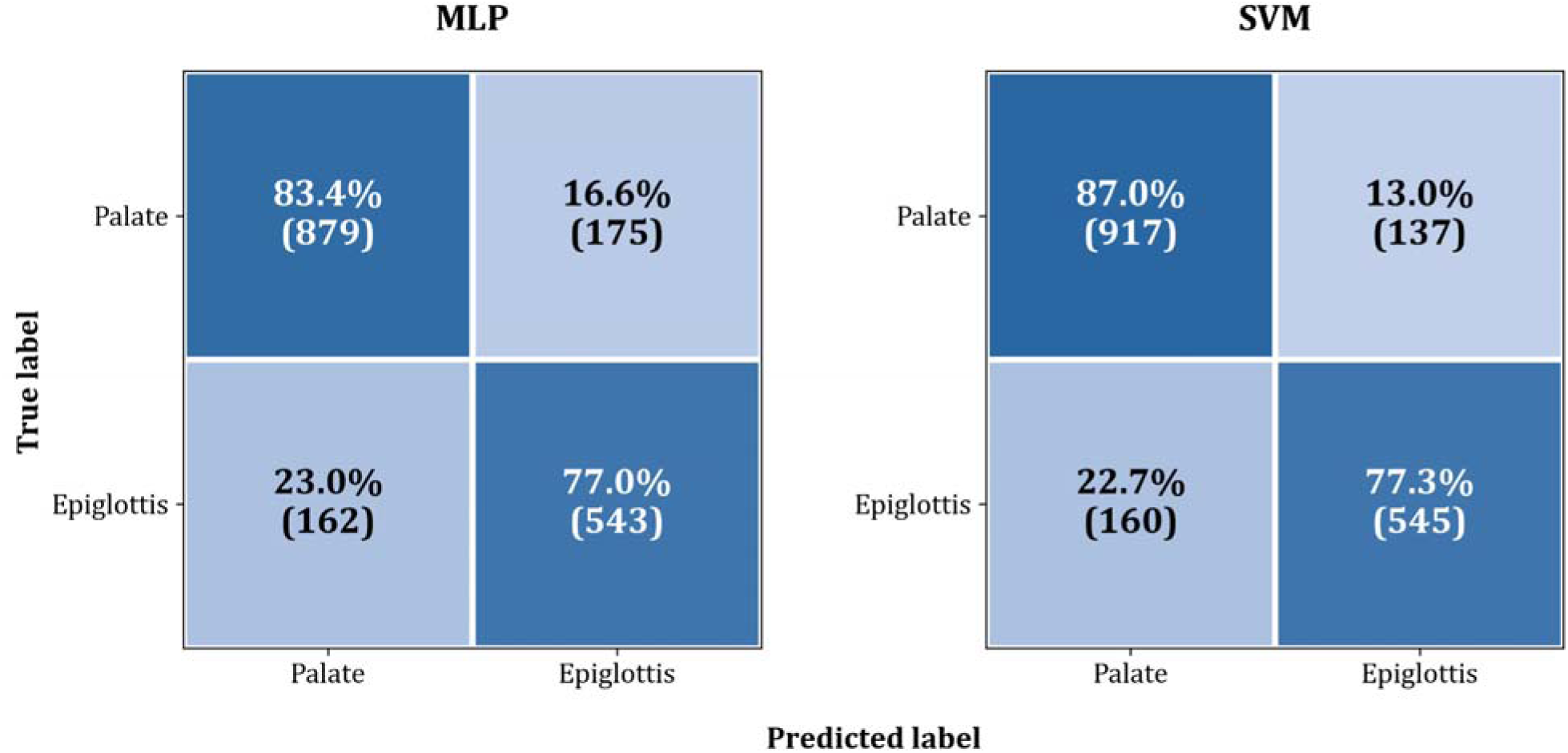
Row-normalized confusion matrices of the classifiers summed across the 5 test folds. Each cell reports the percentage of predictions within the true class (row-wise normalization), with absolute counts in parentheses. The SVM achieved high class-wise recall in identifying palate (87.0%) and good performance on epiglottis (77.3%).

**Table 1.** Summary of Acoustic Features. Summary of all the extracted features divided by domains. A and A^2^ represent the first and second-order derivatives. Statistical descriptors are computed both on the feature and on its derivatives.

| Table 1. Summary of Acoustic Features |  |  |  |
| --- | --- | --- | --- |
| Domain | Feature Group | Statistical Descriptors | N. of Features |
| Cepstral | Mel-Frequency Cepstral Coefficients (Coefficients 1–20, $\Delta$ and $\Delta^2$) | Mean, Median, Std. Dev., Range | 240 |
| Spectral | Flatness and its $\Delta$ and $\Delta^2$ | Mean, Median, Std. Dev., Range | 12 |
| | Centroid and its $\Delta$ and $\Delta^2$ | Mean, Median, Std. Dev., Range | 12 |
| | Bandwidth and its $\Delta$ and $\Delta^2$ | Mean, Median, Std. Dev., Range | 12 |
| | Roll-off (computed using cutoffs 50%, 70% and 90% and their $\Delta$ and $\Delta^2$) | Mean, Median, Std. Dev., Range | 36 |
| | Contrast (of 7 frequency bands and their $\Delta$ and $\Delta^2$) | Mean, Median, Std. Dev., Range | 84 |
| | Chroma features (12 tempered pitches, C to B, and their $\Delta$ and $\Delta^2$) | Mean, Median, Std. Dev., Range | 144 |
|  | Energy distribution of 6 mel bands (20–100 Hz, 100–300 Hz, 300–1000 Hz, 1000–2500 Hz, 2500–5000 Hz, and >5000 Hz) | Mean, Median, Std. Dev., Range | 24 |
| | Flux of 6 frequency bands (20–100 Hz, 100–300 Hz, 300–1000 Hz, 1000–2500 Hz, 2500–5000 Hz, and >5000 Hz )and its $\Delta$ and $\Delta^2$ | Mean, Median, Std. Dev., Range | 84 |
|  | Formants (F1-F4) | Mean, Median, Std. Dev., Range | 16 |
| **Temporal** | Zero-crossing rate and its $\Delta$ and $\Delta^2$ | Mean, Median, Std. Dev., Range | 12 |
|  | Autocorrelation | Mean, Median, Std. Dev., Range, Peak Value, Peak Lag, Peak-to-mean ratio, peak to standard deviation ratio | 8 |
| **Others** | Fundamental Frequency | Voice ratio | 1 |
|  | Vocal Tract Length | Mean, Median, Std. Dev., Range | 4 |
|  | Harmonic-to-noise ratio |  | 1 |
|  | Cepstral Peak Prominence |  | 1 |

**Table 2.** Classification report for MLP across the five test folds (mean and std) in the nested cross-validation. For the palate class: precision 0.85 ± 0.10, recall 0.82 ± 0.08, Fl-score 0.86 ± 0.03. For the epiglottis class: precision 0.79 ± 0.11, recall 0.79 ± 0.10, Fl-score 0.78 ± 0.04. Overall balanced accuracy was 0.83 ± 0.03 and ROC AUC was 0.90 ± 0.02. All these scores were obtained by averaging the results of the classifier models across the five folds and reporting the resulting value together with its standard deviation.

| **Results - MLP** |  |  |  |
| --- | --- | --- | --- |
| **Metric** | **Precision** | **Recall** | **F1-Score** |
| **Palate** | $0.85 \pm 0.10$ | $0.82 \pm 0.08$ | $0.83 \pm 0.06$ |
| **Epiglottis** | $0.75 \pm 0.10$ | $0.78 \pm 0.10$ | $0.76 \pm 0.04$ |
| **Balanced accuracy** | $0.81 \pm 0.03$ | | |
| **ROC AUC** | $0.90 \pm 0.03$ | | |

**Table 3.** Classification report for the SVM across the five test folds (mean and std) in the nested cross-validation. For the palate class: precision 0.85 ± 0.09, recall 0.86 ± 0.07, Fl-score 0.85 ± 0.05. For the epiglottis class: precision 0.79 ± 0.10, recall 0.78 ± 0.08, Fl-score 0.78 ± 0.03. Balanced accuracy was 0.83 ± 0.02 and ROC AUC was 0.90 ± 0.03.

| Results - SVM |  |  |  |
| --- | --- | --- | --- |
| Metric | Precision | Recall | F1-Score |
| Palate | $0.85 \pm 0.09$ | $0.86 \pm 0.07$ | $0.85 \pm 0.05$ |
| Epiglottis | $0.79 \pm 0.10$ | $0.79 \pm 0.08$ | $0.78 \pm 0.03$ |
| Balanced accuracy | $0.83 \pm 0.02$ | | |
| ROC AUC | $0.90 \pm 0.03$ | | |

### Hyperparameter selection

The hyperparameter search spaces for both models are detailed in Tables S1 and S2 (See supplementary materials). Across the five folds the optimal configurations for the SVM consistently identified the RBF kernel with a regularization parameter C = 1 and a kernel coefficient gamma = 0.01. The hyperparameters optimization for the MLP showed stable parameters but a greater variability in finding the optimal architecture: the Adam optimizer, tanh activation, L2 regularization (alpha) of 0.01 and learning rate of 0.001 were quite stable across the 5 folds. Instead, the number of hidden layers ranged from one to four across the 5 folds, with the four-layers architecture being the most selected 2 times out of 5.

### Feature selection results

Following the feature selection procedure, a ranked evaluation was performed to assess the relative predictive value of the retained features across all the 5 outer folds. To improve interpretability, the selected features were grouped into what we call “aggregated features” which encompass all the selected statistics computed for a single acoustic feature (e.g., the mean, median, standard deviation and range of the MFCC1). To quantify the impact of these groups, we computed a score by summing the absolute values of the linear SVM (used within the feature selection) coefficients for all individual statistical descriptors computed for each feature. For instance, the score of Spectral Flux Med-Low is the cumulative importance of its median (selected in 3 folds) and mean (selected in 2 folds). This cumulative metric represents the total predictive contribution of a broader acoustic feature, providing a more robust view of the markers driving the classification than isolated statistical descriptors. The most relevant aggregated features were predominantly related to spectral energy distribution and cepstral characteristics. In particular, spectral flux and overall energy-related features ranked highest in terms of importance. Mel-Frequency Cepstral Coefficients (MFCCs), spanning multiple coefficients, also showed a strong contribution. Additionally, spectral contrast-based features and temporal dynamics (delta features) were consistently represented among the top-ranked predictors. The top 10 aggregated features are shown in Figure 2 and their relative composition, and aggregated scores are provided in Table 4.

**Figure 2:**
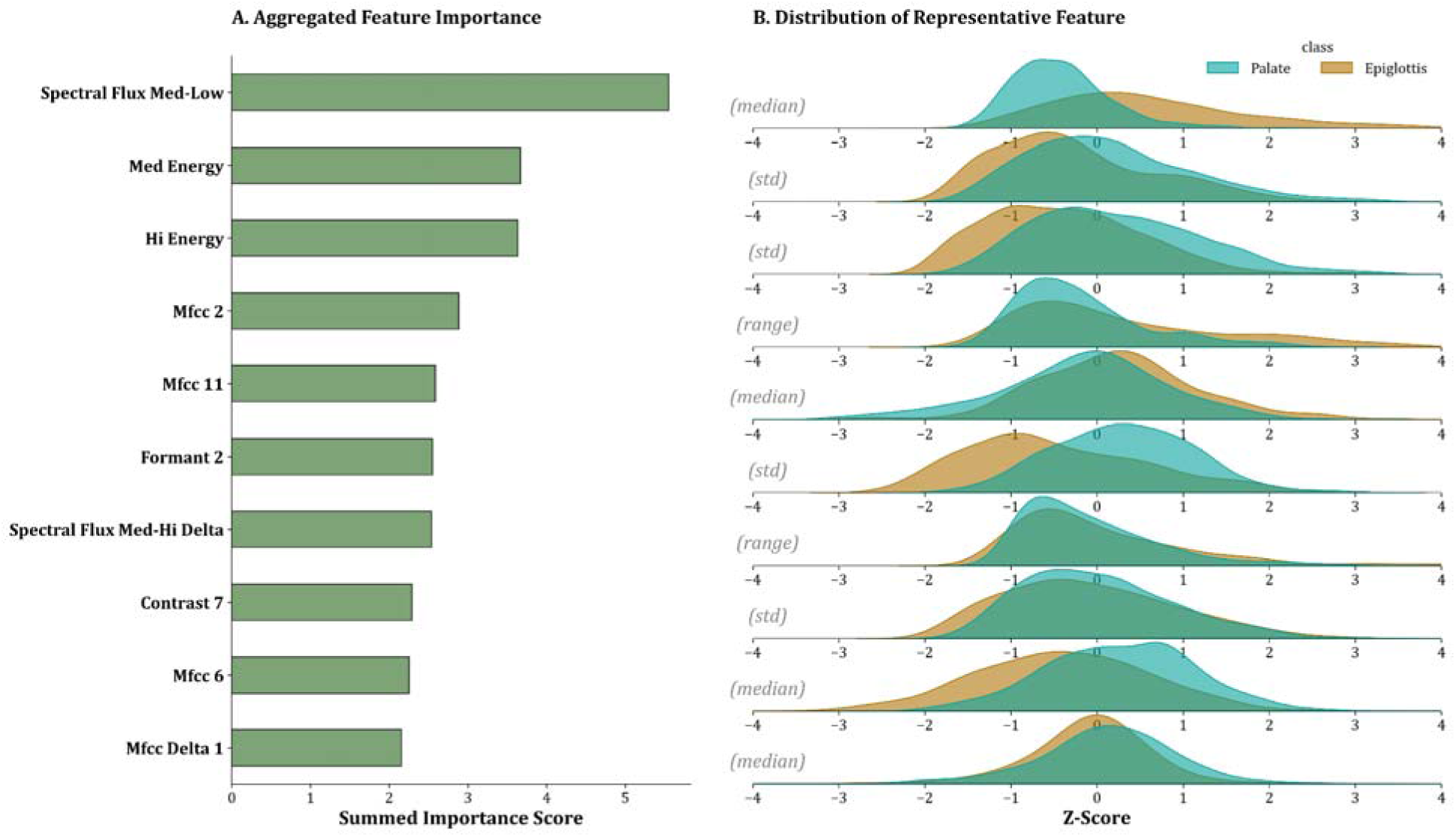
**A:** Aggregated feature importance scores contributing to the model (the linear SVM trained on the post-pruning feature set, with feature importance computed from model coefficients at each iteration of RFE) across 5 cross-validation folds. The total score for each aggregated feature is the sum of the descriptive statistics (mean. Median, standard deviation, range) of a feature. The frequency bands were defined as following: 20-100 Hz = sub-band (or sub), 100-300 Hz = low, 300-1000 Hz = medium- low (or med-low), 1000-2500 Hz = medium (or med), 2500-5000 Hz = medium-high (or med-hi), and >5000 Hz = high (or hi). B: Standardized distributions (Z-scores) of the most representative feature for each aggregated feature, identified as the primary contributor to the global score. In the case of Spectral Flux Med-Low, its median is shown as the representative distribution for the Palate and Epiglottis classes, as it has been selected as the best feature in most of the folds.

**Table 4.** Top-10 ranked feature aggregations contributing to the model (the linear SVM trained on the post-pruning feature set, with feature importance computed from model coefficients at each iteration of RFE) across 5 cross validation folds. The frequency bands were defined as following: 20-100 Hz = sub-band (or sub), 100-300 Hz = low, 300-1000 Hz = medium-low (or med-low), 1000-2500 Hz = medium (or med), 2500-5000 Hz = medium-high (or med-hi), and >5000 Hz = high (or hi). The column Composition specifies the statistical descriptors for each feature. The number inside the round brackets close to the statistical descriptor indicates the occurrences of the individual feature across the 5 folds. The aggregation score is computed by summing the absolute values of the linear SVM coefficients for all individual features belonging to each aggregation. Spectral flux (med/low) shows the highest importance, followed by medium and high energy features, while several MFCC components and formant-based measures provide additional but comparatively smaller contributions.

| Top 10 Best aggregated features |  |  |
| --- | --- | --- |
| Aggregated features | Composition | Aggregated feature Score |
| Spectral flux med/low | Median (3), mean (2) | 5.55 |
| Medium energy | Standard deviation (4) | 3.66 |
| Hi energy | Median (1), standard deviation (5) | 3.63 |
| MFCC 2 | Range (4), median (1) | 2.88 |
| MFCC 11 | Median (4) | 2.58 |
| Formant 2 | Standard deviation (3), mean (1) | 2.55 |
| Spectral Flux med/hi delta | Standard deviation(1) , range (3) | 2.53 |
| Contrast 7 | Standard deviation (3) | 2.29 |
| MFCC 6 | Median (2), range (2) | 2.25 |
| MFCC delta 1 | Median (4) | 2.15 |

## Discussion

In the present study the ability of a machine learning algorithm in correctly classify snoring events in palatal or epiglottic on the sole base of the sound produced during snoring was evaluated. The results here reported suggest that machine learning can accurately discriminate between palatal and epiglottic snoring using acoustic information alone. Using high-confidence event-level anatomical labels obtained through direct endoscopic visualization during Drug-Induced Sleep Endoscopy (DISE) and a rigorous patient-independent nested cross-validation framework, both evaluated classifiers achieved robust discrimination (ROC AUC ∼ 0.90), supporting the hypothesis that snoring contains clinically meaningful information reflecting the anatomical site of upper airway obstruction. Importantly, these results were obtained despite class imbalance and a strictly patient-independent validation strategy, conditions that more closely resemble real-world clinical deployment than conventional segment-based validation approaches. Rather than representing merely a symptom of OSA, snoring appears to encode reproducible biomechanical signatures that can be exploited for non-invasive anatomical phenotyping through artificial intelligence.

The concept that snoring carries anatomical information is supported by decades of physiological research. In their landmark review, Pevernagie et al. (25) described snoring as the consequence of a complex interaction between inspiratory airflow and compliant upper-airway tissues, demonstrating how tissue mass, stiffness, geometry and airflow dynamics determine its acoustic properties. Accordingly, different anatomical structures are expected to generate distinct acoustic signatures rather than isolated frequency peaks. Clinical studies progressively confirmed this concept. Miyazaki et al. (26) first demonstrated an association between snoring frequency and the anatomical level of obstruction, while Lee et al. (27) and Qualickuz Zanan et al. (28) further showed that snoring acoustics correlate with DISE findings and may even predict surgical outcomes. Nevertheless, considerable heterogeneity in recording protocols, patient populations and reference standards has so far limited the clinical translation of these observations.

Recent advances in artificial intelligence have substantially expanded the diagnostic potential of snoring analysis. Rather than relying on predefined frequency thresholds, machine learning integrates multiple complementary acoustic descriptors to identify multidimensional patterns associated with upper-airway obstruction. However, recent reviews have consistently highlighted important methodological limitations. Huang et al. (29) emphasized the marked heterogeneity of available studies, whereas the systematic review by Tartaglia et al. (30) identified anatomical obstruction-site classification as one of the most promising emerging applications of artificial intelligence while simultaneously stressing the need for larger clinically annotated datasets, standardized methodologies and rigorous validation strategies.

Although previous studies have successfully demonstrated the feasibility of machine learning-based obstruction-site classification (31,32), our study was intentionally designed from a different perspective. Rather than pursuing comprehensive VOTE classification or maximizing algorithmic performance, we addressed a focused clinical question: can acoustic analysis reliably distinguish between the two obstruction sites with the greatest immediate therapeutic implications? This strategy was not dictated by computational convenience but by clinical relevance. Expanding classification from two to four anatomical classes inevitably requires substantially larger and well-balanced datasets, particularly for relatively underrepresented phenotypes such as epiglottic collapse. We therefore deliberately adopted a stepwise strategy, aiming first to establish a robust and clinically reliable binary classifier before progressively extending the framework to additional obstruction sites.

This approach also enabled the creation of one of the largest event-level datasets specifically dedicated to palatal and epiglottic snoring currently reported in the literature. Unlike studies assigning a single obstruction pattern to an entire patient or overnight recording, every snoring event in our dataset was independently annotated by two blinded expert examiners according to the anatomical structure actively generating the sound, and only events demonstrating complete inter-observer agreement were retained. This strategy minimized temporal ambiguity, reduced annotation bias and provided a high-confidence anatomical ground truth for supervised machine learning. Furthermore, patient-independent nested cross-validation and feature selection performed exclusively within each training fold minimized information leakage and optimistic bias, providing a realistic estimate of model performance in previously unseen subjects.

An additional strength of the proposed framework lies in the physiological interpretability of the selected acoustic features. Despite the initial extraction of 691 descriptors, recursive feature elimination consistently converged towards a relatively small subset dominated by spectral features, particularly MFCCs, spectral energy distribution, spectral contrast and spectral flatness. Notably, the highest-ranking variables predominantly described energy distribution within the medium-frequency range (approximately 300-1000 Hz), suggesting that this spectral region carries the greatest anatomical information for distinguishing palatal from epiglottic collapse. These findings are physiologically plausible, as snoring originates from the interaction between airflow dynamics and the biomechanical properties of compliant upper-airway tissues, resulting in complex spectral fingerprints rather than isolated frequency components (25). Interestingly, variables related to the fundamental frequency (F0) consistently failed to survive feature selection. Unlike speech, snoring is primarily generated by irregular tissue vibration and turbulent airflow rather than periodic vocal-fold oscillation, making multidimensional spectral descriptors considerably more informative than pitch-related measures. The comparable performance achieved by both SVM and MLP further supports this interpretation, suggesting that successful anatomical phenotyping depends more on the quality of anatomical annotation and feature representation than on algorithmic complexity itself.

The clinical value of the proposed approach extends beyond the development of an accurate classification model. Our objective is not to replace Drug-Induced Sleep Endoscopy, but to make its use more efficient. DISE remains the reference standard for dynamic evaluation of upper-airway collapse; however, its cost, invasiveness and limited availability prevent its routine use in many centres. We therefore envision acoustic analysis as a decision-support tool capable of improving patient selection and facilitating anatomical phenotyping before endoscopic evaluation.

The decision to focus specifically on palatal and epiglottic obstruction reflects this clinical philosophy. These two anatomical phenotypes represent the principal surgical targets in contemporary sleep surgery but differ substantially in their therapeutic implications. While isolated palatal obstruction may respond to several treatment modalities—including positive airway pressure, mandibular advancement devices, myofunctional therapy, hypoglossal nerve stimulation and different palatal surgical procedures—primary epiglottic collapse frequently shows limited response to conservative therapies and often requires dedicated surgical correction (33). Consequently, distinguishing these two obstruction patterns is not merely an anatomical classification task but a clinically actionable decision capable of directly influencing treatment planning.

Rather than serving as a definitive diagnostic tool, the proposed model should therefore be considered a first-line anatomical screening instrument. Patients predicted to have predominant epiglottic collapse could be prioritized for referral to dedicated sleep surgery centres, while DISE would retain its central role in confirming the anatomical phenotype, defining collapse configuration and planning treatment. Similarly, in patients with failure of first-line therapies such as CPAP or mandibular advancement devices, pre-endoscopic identification of a possible epiglottic collapse could increase clinical suspicion and facilitate interpretation of subsequent DISE findings. Although these applications remain speculative and require prospective validation, they illustrate how artificial intelligence may improve the efficiency of existing diagnostic pathways rather than replace established investigations.

The present study has several methodological strengths. High-confidence anatomical labels were obtained through independent double-blinded expert annotation with inclusion restricted to events demonstrating complete agreement, ensuring reliable event-level ground truth. The study design further minimized methodological bias through patient-independent nested cross-validation and feature selection performed exclusively within the training folds. Finally, the use of established and inherently interpretable machine learning algorithms shifted the focus from algorithmic novelty to clinical applicability, allowing direct physiological interpretation of the acoustic features contributing to classification.

Nevertheless, several limitations should be acknowledged. This was a retrospective single-centre study, and all recordings were acquired using a standardized protocol. Although independent blinded annotation strengthened the reliability of the reference standard, external validation across different institutions, recording systems and patient populations remains essential before routine clinical implementation. Furthermore, the study population was intentionally restricted to normal-weight patients with moderate, non-positional OSA to minimize potential confounding factors related to obesity-associated airway collapse and positional dependency. While this homogeneous population strengthened the internal validity of the study, it may limit the generalizability of the proposed framework to the broader OSA population.

Restricting the model to two anatomical classes should also be interpreted as a deliberate methodological strategy rather than a technical limitation. Reliable multiclass machine learning requires adequate representation of every anatomical phenotype, and increasing the number of output classes inevitably demands substantially larger annotated datasets. We therefore intentionally prioritized the development of a robust binary classifier addressing a clinically relevant question before progressively extending the framework towards comprehensive anatomical phenotyping.

Future research should primarily focus on expanding clinically annotated datasets through multicenter collaboration rather than increasing algorithmic complexity. We believe that the next major challenge is the creation of large, standardized databases combining synchronized DISE recordings and snoring acoustics, allowing robust representation of additional obstruction sites and improving external validity. Such collaborative efforts will facilitate the progressive development of reliable multiclass models capable of reflecting the complexity of upper-airway obstruction encountered in routine clinical practice.

### Conclusions

In conclusion, the present study demonstrates that snoring is not merely a symptom of obstructive sleep apnea but a clinically informative acoustic biomarker capable of reflecting upper-airway anatomy. Using high-confidence DISE-derived anatomical labels, rigorous patient-independent validation and established machine learning techniques, we demonstrate that palatal and epiglottic obstruction can be reliably distinguished using acoustic information alone. This work proposes a clinically oriented framework for non-invasive anatomical phenotyping and establishes the basis for future multicenter development toward comprehensive acoustic characterization of upper-airway obstruction.

## Data Availability

All data produced in the present study are available upon reasonable request to the authors

## Author Contributions

Conceptualization: Francesco Mozzanica, Federico Leone.

Methodology: Francesco Pietrogiacomi, Emanuele Agrimi, Linda Fiorini.

Investigation: All authors.

Formal analysis: Francesco Pietrogiacomi, Emanuele Agrimi, Linda Fiorini.

Data curation: Francesco Pietrogiacomi, Emanuele Agrimi, Linda Fiorini, Valerio Cesarini, Federica Vultaggio.

Visualization: Francesco Pietrogiacomi, Linda Fiorini, Emanuele Agrimi.

Writing - original draft preparation: Francesco Pietrogiacomi, Federico Leone, Francesco Mozzanica, Emanuele Agrimi, Linda Fiorini, Giorgio Gnecco.

Writing - review and editing: All authors.

Supervision: Francesco Mozzanica, Federico Leone, Giorgio Gnecco.

Project administration: Federico Leone, Francesco Mozzanica.

Funding acquisition: Francesco Mozzanica.

All authors have read and agreed to the published version of the manuscript.

## Competing Interests

The authors declare no competing interests.

## Appendix

## Acknowledgements

This project was partially funded by IRCCS Multimedica through the grant: NEW IDEAS AWARD 2024.

## Notes

### Competing Interest Statement

The authors have declared no competing interest.

### Author Declarations

Ethics committee/IRB of Istituto Auxologico Italiano IRCCS gave ethical approval for this work (Protocol No. 4150-ID).

