## Supplement Materials for "The sound of snoring: AI-based acoustic phenotyping of upper airway obstruction in Obstructive Sleep Apnea"

for

**Affiliations**:

### *Data processing*

Following extraction and manual trimming, the audio samples underwent volume normalization using the Root Mean Square (RMS) normalization to account for variations in microphone distance, sensitivity, and patient movement. The RMS value of a discrete signal *x[n]* of length *N* was computed for each audio clip as:

$$RMS = \sqrt{\frac{1}{N}{\sum_{n=1}^{N} x[n]^{2}}}$$

The audio samples were then scaled so that each clip had the same RMS value, using the lowest as a target in order to avoid positive gain that could lead to clipping, in turn ensuring that the distribution of signal amplitudes was standardized across all recordings. Finally, all the clips were resampled from 44,100 Hz to 22,050 Hz to optimize computational efficiency and ensure a uniform frequency range for subsequent feature extraction. This choice was made to reduce the number of samples for computational efficiency while taking into account that snoring-like sounds contain most of their relevant information at lower frequencies [1]. Moreover, the selected downsampling rate still preserves frequency content until 10 kHz. These preprocessing steps ensured the reliability and comparability of the acoustic data for the classification tasks.

### *Feature Extraction*

A comprehensive set of cepstral, spectral, temporal, and harmonic descriptors was extracted from each preprocessed audio segment. For dynamic descriptors, the mean, median, standard deviation, and range were computed whenever applicable. First- and second-order temporal derivatives (Δ and Δ²) were also calculated for selected features. The complete procedure yielded 691 features per snoring event.

Cepstral descriptors included the following:

- **Mel-Frequency Cepstral Coefficients** (**MFCC**) [2], which quantify the presence of cyclical events in the auditory spectrum, were computed (MFCCs 1–20), along with their first- and second-order derivatives (Δ and Δ²). MFCCs are vectors derived from a Fourier-like (Discrete Cosine) transform applied to the frequency-domain representation of the signal, after logarithmic weighting and division of the spectrum into 20 mel-bands, reshaping the frequency axis to be equally spaced from a perceptual point of view. For each coefficient, the following statistical descriptors were computed: mean, std.dev. (standard deviation), median, and range, yielding a total of 240 MFCC-based features.

Spectral descriptors included the following:

- **Spectral flatness** [3] (mean, std.dev., median, range and its Δ and Δ² components), which estimates the noise-like quality of a spectrum by computing its “flatness” as in peak-less nature;
- **Spectral centroid** [4] (mean, std.dev., median, range and its Δ and Δ² components), which is an estimate of the center of mass of the spectral curve computed via weighted mean of the frequencies, and is related to perceptual presence/absence of “brightness”;
- **Spectral bandwidth** [5] (mean, std.dev., median, range and its Δ and Δ² components), as the spread of frequencies around the centroid;
- **Spectral roll-off** [6] (mean, std.dev., median, range and its Δ and Δ² components), which indicates the frequency below which a given percentage of the signal energy is contained, computed at three cutoffs: 50%, 70% and 90%;
- **Spectral contrast** (mean, std.dev., median, range of the function and its Δ and Δ² components) across seven frequency bands, quantifying the difference of energy between the peak region (top 20%) and lower region (bottom 20%);
- **Chroma features** [7] (mean, std.dev., median, range of the function and its Δ and Δ² components), representing the distribution of energy across the 12 tempered pitch classes (musical notes from C to B, where A = 440 Hz);
- **Energy distribution** (mean, standard deviation, median, and range) across six mel-frequency bands: 20–100 Hz, 100–300 Hz, 300–1000 Hz, 1000–2500 Hz, 2500–5000 Hz, and >5000 Hz; In the following, these bands we selected are called respectively sub-band (or sub), low, medium-low (or med-low), medium (or med), medium-high (or med-hi), high (or hi);
- **Spectral flux** [8] (mean, std.dev., median, range of the function and its Δ and Δ² components), which entails the evolution of the spectrum in time (spectrogram) and compares the power spectrum for one frame against that from the previous frame, computed for the whole spectrum and also for the six mel bands;
- **Formants** (mean, std.dev., median, and range for F1–F4), estimated via Linear Predictive Coding (LPC). They are defined as the relevant peaks in the smooth envelope of the spectrum and are known to be related to sound openness/closeness as well as vocal-like sounds, and are dependent on the physical properties of the vocal tract.

Temporal features included the following:

- **Zero-crossing rate** (mean, std.dev., median, range of the function and its Δ and Δ² components), representing how frequently the signal waveform crosses zero (an indicator of signal noisiness or high-frequency content);
- **Autocorrelation-based measures**: mean, standard deviation, median, range, peak value, peak lag, peak-to-mean ratio, and peak-to-standard deviation ratio.

Additional features included:

- **Vocal Tract Length** (**VTL**) (mean, std.dev., median, range). For each frame, independent length estimates *L* were derived from the first *n* formant frequencies $F_{n}$ as shown using the formula $L= \frac{(2n - 1) \cdot c}{4 \cdot F_{n}}$ , where *c* is the speed of sound (35,000 cm/s). These individual estimates were then averaged within each frame to provide a stable VTL value [9];
- **Harmonics-to-Noise Ratio** (**HNR**) (mean, std.dev., median, range, Δ, and Δ²) quantifies the ratio between periodic and aperiodic components. It focuses on the temporal stability (consistency between cycles) of the signal; lower values indicate vibratory instability and irregular airway dynamics, while higher values indicate harmonic and periodic stability;
- **Cepstral Peak Prominence** (**CPP**) (mean, std.dev., median, range, Δ, and Δ²) measures the magnitude of the highest cepstral peak relative to its regression line (noise floor). Since Cepstrum is a measure of the clearness of spectral peaks (namely F0 and formants), a high peak prominence indicates a highly intelligible fundamental frequency and a less noise-like spectrum;
- **F0** (**voiced/non voiced ratio**), which is defined as the main vibrational component in a non-noisy signal, and is related to the perceived pitch. F0 is computed as an estimate based on the principle that the main frequency in a periodic signal creates the first local maximum after t=0 in the autocorrelation function. The algorithm we chose is YIN [10], which also outputs NaN (not-a-number) values when no suitable F0 candidates are found. This was especially useful given the noise-like nature of the analyzed sounds. The YIN algorithm was applied to each audio segment, thus creating a vector of F0 measures for each single event. From the F0 vector, the voicing ratio was derived. This metric represents the proportion of the snoring event characterized by periodic vibration, calculated as the ratio of frames with a detected F0 to the total number of frames in the segment.

This feature extraction process yielded a final set of 691 features for each audio segment. For a graphical summary of the entire set of the extracted features, see Table S0. Feature extraction focused primarily on spectral descriptors, given their proven utility in previous studies on similar tasks [11–13]. Additionally, F0 was analyzed to test the hypothesis that different site types may produce systematically different pitch patterns.

Indeed, despite snoring being a mainly non-vocalised sound, we chose to also analyze the presence of fundamental frequency (F0) within each segment. Vocalised sounds yield a clear, definite F0 that evolves through speech, whereas sounds like snoring may entail one of the following possibilities: 1) There may be a superimposed vibration on the vocal folds which in turns created a pseudo-vocalised sound alongside the main snoring; 2) The snoring itself may yield a vibration with a definite F0; 3) Snoring may contain noise-like components that do not yield a definite F0.

*Classification Model and Training Strategies*

Two machine learning models were implemented for the binary classification of airway obstruction as palatal or epiglottic. The first model was a Support Vector Machine (SVM) [14], chosen for its efficiency in high-dimensional spaces and its ability to define optimal decision boundaries. The second was a Multi-Layer Perceptron (MLP) [15], a feedforward neural network designed to capture complex non-linear relationships. To ensure robust model generalizability and prevent optimistic bias, a 5x5 nested cross-validation (NCV) procedure was employed [16]. The outer loop was used for unbiased performance evaluation, while the inner loop (embedded within each outer training fold) was dedicated to hyperparameter optimization via grid search. For each outer fold, the training data was first standardized using a StandardScaler, followed by the identification of the optimal feature subset (see Section: *Feature Selection*). Using these selected features, the inner loop explored various kernels and regularization parameters for the SVM, and multiple hidden layer architectures, activation functions, and regularization parameters for the MLP; the hyperparameter space for both models is detailed in Tables S1 and S2 (See supplementary materials). To address class imbalance, strategies were tailored to each model: for the SVM, a sample weighting strategy was applied to balance the contribution of each subject and class; for the MLP, a random oversampling strategy was employed to ensure an equal distribution of classes during training. Once the optimal hyperparameters were identified within the inner loop, the models were retrained on the full training set of the current outer fold–using the previously selected 20 features–before being evaluated on the held-out test set. Two critical constraints ensured clinical validity: 1) Patient-wise splitting: to prevent data leakage, a patient-based splitting strategy was strictly maintained across both the inner and outer loops. All segments from a given patient were confined exclusively to either the training or the testing/validation sets, ensuring that the models were always optimized and evaluated on entirely 'unseen' clinical populations rather than patient-specific acoustic signatures; 2) Class Stratification: while strictly adhering to the subject-wise constraint, stratification was enforced to ensure that the training and test sets in the outer loop maintained a class distribution representative of the overall dataset. This ensured that each fold remained balanced and representative of the clinical population under study; the complete distribution of subjects and snoring segments across both classes is detailed in Table S3. Performance was evaluated on the held-out test sets using balanced accuracy as the primary metric, alongside precision, recall, F1-score, and the Area Under the Receiver Operating Characteristic curve (AUC-ROC).

### *Feature Selection*

To avoid data leakage, feature selection was performed independently within each outer fold of the cross-validation procedure, ensuring the selection process was never influenced by the test data. The procedure followed two sequential steps: 1) Multicollinearity Pruning: Redundancy was reduced by computing pairwise Pearson correlation coefficients (*r*) for all features. When *r > 0.80*, the features were considered highly collinear. To resolve these ties, the point-biserial correlation between each feature and the clinical target was computed; only the feature with the higher correlation to the target was retained; 2) Recursive Feature Elimination [17]**:** The pruned set was further refined using RFE with a linear SVM as the base estimator. This algorithm iteratively assessed feature importance based on the model’s coefficients, removing the least informative 10% of features at each step. This process continued until a final subset of 20 features was reached.

| **Table 1. Summary of Acoustic Features** | | | |
| --- | --- | --- | --- |
| **Domain** | **Feature Group** | **Statistical Descriptors** | **N. of Features** |
| **Cepstral** | Mel-Frequency Cepstral Coefficients (Coefficients 1–20, Δ and Δ²) | Mean, Median, Std. Dev., Range | 240 |
| **Spectral** | Flatness and its Δ and Δ² | Mean, Median, Std. Dev., Range | 12 |
|  | Centroid and its Δ and Δ² | Mean, Median, Std. Dev., Range | 12 |
|  | Bandwidth and its Δ and Δ² | Mean, Median, Std. Dev., Range | 12 |
|  | Roll-off (computed using cutoffs 50%, 70% and 90% and their Δ and Δ²) | Mean, Median, Std. Dev., Range | 36 |
|  | Contrast (of 7 frequency bands and their Δ and Δ²) | Mean, Median, Std. Dev., Range | 84 |
|  | Chroma features (12 tempered pitches, C to B, and their Δ and Δ²) | Mean, Median, Std. Dev., Range | 144 |
|  | Energy distribution of 6 mel bands (20–100 Hz, 100–300 Hz, 300–1000 Hz, 1000–2500 Hz, 2500–5000 Hz, and >5000 Hz) | Mean, Median, Std. Dev., Range | 24 |
|  | Flux of 6 frequency bands (20–100 Hz, 100–300 Hz, 300–1000 Hz, 1000–2500 Hz, 2500–5000 Hz, and >5000 Hz )and its Δ and Δ² | Mean, Median, Std. Dev., Range | 84 |
|  | Formants (F1-F4) | Mean, Median, Std. Dev., Range | 16 |
| **Temporal** | Zero-crossing rate and its Δ and Δ² | Mean, Median, Std. Dev., Range | 12 |
|  | Autocorrelation | Mean, Median, Std. Dev., Range, Peak Value, Peak Lag, Peak-to-mean ratio, peak to standard deviation ratio | 8 |
| **Others** | Fundamental Frequency | Voice ratio | 1 |
|  | Vocal Tract Length | Mean, Median, Std. Dev., Range | 4 |
|  | Harmonic-to-noise ratio |  | 1 |
|  | Cepstral Peak Prominence |  | 1 |

**Table S1**. Summary of extracted acoustic features. Note. Δ and Δ² denote first- and second-order derivatives. Statistical descriptors were calculated for the feature and, where applicable, for its derivatives.

| **Hyperparameter Space - SVM** | | |
| --- | --- | --- |
| **Kernel** | **Hyperparameters** | **Values** |
| linear | *C* | 0.1, 1, 10, 35, 50 |
| **rbf** | *C* | 0.01, 0.1, **1**, 10, 50, 100, 150, 200 |
|  | γ | scale, **0.01**, 0.1, 1 |
| poly | *C* | 0.05, 0.1, 1, 10 |
|  | γ | scale, auto |
|  | *R* | 0, 1 |
|  | Degree | 2,3 |

**Table S2**: Hyperparameter space of the SVM explored via grid search. **Bold values** were the most selected across the 5 folds of the outer loop. The kernel rbf γ = 0.01 wer selected 4 times. *C* value was 1 for all the folds.

| **Hyperparameter Space - MLP** | |
| --- | --- |
| **Hyperparameters** | **Values** |
| Hidden Layers Sizes | (100) (150) (100, 50) (150, 100, 50) **(200, 150, 75, 50)** |
| Alpha | 0.1, **0.01**, 0.001, 0.0001 |
| Learning rate | 0.1, 0.01, **0.001** |
| Solver | **adam**, lbfgs |
| Activation function | **tanh**, relu |

**Table S3**: Hyperparameter space of the MLP explored via nested grid search. Values for *Hidden Layer Sizes* specify the network architecture as (neurons_layer_1, neurons_layer_2, ...); for instance, (100, 50) denotes a 2-layer architecture with 100 and 50 neurons in the first and second hidden layers, respectively. **Bold values** indicate the hyperparameter choices selected most frequently across the 5 outer cross-validation folds. Specifically, the (200, 150, 75, 50) hidden layer architecture was selected twice, alpha = 0.01 three times, an initial learning rate of 0.001 three times, and the tanh activation function four times. The adam solver was chosen as the optimization algorithm across all 5 folds..

| **Distribution of samples across 5 folds** | | | | | |
| --- | --- | --- | --- | --- | --- |
| **FOLD** | **SET** | **SAMPLES** | **SUBJECTS** | **PALATE** | **EPIGLOTTIS** |
| **1** | TRAIN | 1324 | 127 | 783 | 541 |
|  | TEST | 435 | 32 | 271 | 164 |
| **2** | TRAIN | 1419 | 128 | 877 | 542 |
|  | TEST | 340 | 31 | 177 | 163 |
| **3** | TRAIN | 1420 | 127 | 781 | 639 |
|  | TEST | 339 | 32 | 273 | 66 |
| **4** | TRAIN | 1465 | 126 | 868 | 597 |
|  | TEST | 294 | 33 | 186 | 108 |
| **5** | TRAIN | 1408 | 128 | 907 | 501 |
|  | TEST | 351 | 31 | 147 | 204 |

**Table S4**. Sample and subject partitioning across 5 outer cross-validation folds (N = 1,759). Values indicate the number of total samples, subjects, and target class distributions (Palate vs. Epiglottis) for training and test sets per fold. To prevent data leakage, a stratified group split was applied at the subject level, ensuring that segments from the same subject were assigned exclusively to either the training or test set within any fold. Every subject was included in the test set exactly once across the 5 folds. Model hyperparameters were tuned via inner cross-validation within each training set before evaluating performance on the held-out test set.
